# Genetic background of grey matter brain networks related to attention-deficit/hyperactivity disorder

**DOI:** 10.1101/2022.01.07.22268852

**Authors:** Gido H. Schoenmacker, Kuaikuai Duan, Kelly Rootes-Murdy, Wenhao Jiang, Pieter J. Hoekstra, Catharina A. Hartman, Jaap Oosterlaan, Martine Hoogman, Barbara Franke, Jessica A. Turner, Jingyu Liu, Tom Claassen, Tom Heskes, Jan K. Buitelaar, Alejandro Arias Vásquez

## Abstract

Attention-deficit/hyperactivity disorder (ADHD) is a common neurodevelopmental disorder and is associated with structural grey matter differences in the brain. We investigated the genetic background of some of these brain differences in a sample of 899 adults and adolescents consisting of individuals with ADHD and healthy controls. Previous work in an overlapping sample identified three ADHD-related grey matter brain networks located in areas of the superior, middle, and inferior frontal gyrus as well as the cerebellar tonsil and culmen. We associated these brain networks with protein coding genes using a statistical stability selection approach. We identified ten genes, the most promising of which were NR3C2, TRHDE, SCFD1, GNAO1, and UNC5D. These genes are expressed in brain and linked to neuropsychiatric disorders including ADHD. With our results we aid in the growing understanding of the aetiology of ADHD from genes to brain to behaviour.

## Introduction

Attention-deficit/hyperactivity disorder (ADHD) is a common neurodevelopmental disorder, affecting an estimated 3.4-7.2% of children (Polanczyk *et al*., 2007, 2014; Thomas *et al*., 2015) and 2-3% of adults (Simon *et al*., 2009; Fayyad *et al*., 2017) worldwide. ADHD is characterised by symptoms in the domains of inattention and/or hyperactivity-impulsivity and resulting impairment of functioning (American Psychiatric Association, 2013) and is heterogeneous in aetiology, developmental trajectory, and clinical profile (Luo *et al*., 2019). In 60-80% of cases, ADHD presents with comorbid psychiatric disorders such as oppositional defiant and conduct disorders, autism spectrum disorder, depression, bipolar disorder, borderline personality disorder, anxiety disorders, as well as substance use disorders, sleep disorders, and learning disabilities (Jensen *et al*., 1997; Ohnishi *et al*., 2019; Kooij *et al*., 2012; Katzman *et al*., 2017; Biederman *et al*., 1993).

Twin studies have shown that childhood ADHD is highly heritable with genetic factors explaining over 70% of variance (Faraone *et al*., 2015). For ADHD in adults, heritability is lower than or comparable to childhood ADHD, with self-ratings showing lower heritability (<50%) than ratings by others (Faraone and Larsson, 2019). Despite this high heritability, the search for genetic factors underlying ADHD has proven difficult (Faraone *et al*., 2005). Only the largest genome-wide association (GWA) meta-analysis to date reported genome-wide significant loci, concluding that common genetic variants account for 22% of phenotypical variance (Demontis *et al*., 2019). The polygenic nature of ADHD means that individual genetic effects are small, with odds ratios between 1.077-1.198. These is also evidence for a shared genetic background of childhood and adult ADHD (Rovira *et al*., 2020). In addition, there is considerable overlap between the genetics of ADHD and neurodevelopmental traits in the general population (Martin *et al*., 2014) as well as other psychiatric disorders (Anttila *et al*., 2018) such as autism spectrum disorder (Ghirardi *et al*., 2019; Stergiakouli *et al*., 2017).

Since ADHD is at least in part a disorder of the brain, it may be expected that the influence of genetic variants on ADHD is mediated by variations in brain structure and function. Indeed, differences in brain development, volume, activity, and connectivity are observed between groups of individuals with ADHD and healthy controls (Klein *et al*., 2017; Hoogman *et al*., 2017, 2019). Using magnetic resonance imaging (MRI), specific structural differences in grey matter (GM) tissue have been identified. On average, individuals with ADHD have 3-5% overall reduction in GM volume (Greven *et al*., 2015; Hoogman *et al*., 2019). More specifically, ADHD status is associated with reduced cortical surface and thickness (Hoogman *et al*., 2019) as well as reduction in subcortical structures including nucleus accumbens, amygdala, caudate nucleus, hippocampus, and putamen (Hoogman *et al*., 2017). These case-control differences were most pronounced in children. In addition, ADHD in the general population has been associated with reductions in total brain volume (Hoogman *et al*., 2012; Shaw *et al*., 2011) as well as GM volume reductions in frontal, temporal, caudate nucleus, and cerebellar regions (Xavier Castellanos *et al*., 2002).

Studies combining neuroimaging and genetics approaches in ADHD face all the challenges of a genetically and biologically heterogeneous disorder with small effect sizes, complicated by overall small sizes of samples due to the need for both neuroimaging and genetic information to be available. ADHD imaging genetics studies mostly focussing on single genetic variants have so far produced few promising results (Klein *et al*., 2017). Recently, a multivariate imaging genetic analysis approach, named sparse parallel independent component analysis, has been developed to optimize the independence and sparsity of genetic sources as well as the imaging-genetic associations, and it has been applied to identity the genetic factors underlying GM volume alterations in superior frontal gyrus associated with working memory underperformance in both ADHD adults and adolescents (Duan *et al*., 2020). Meanwhile, the first significant genome-wide genetic results on the genetic covariation between ADHD and intracranial volume were reported by combining large (n=11,221-55,374) GWA meta analyses, thereby circumventing the need for both imaging and genetic data to be available for the same subjects (Klein *et al*., 2019). In contrast, in the present study we worked with raw imaging and genetics data of 899 individuals and addressed the statistical power issues differently.

Specifically, we engaged ADHD imaging genetics power issues on three fronts. Firstly, we used previously published brain networks associated with ADHD (Liu *et al*., 2020; Duan *et al*., 2018). By looking at the most discriminative combinations of brain volumes instead of fixed structural regions, we increased our power to detect ADHD-related differences by limiting the number of comparisons (Xu *et al*., 2009). Secondly, we reduced information at the genetic level by running a gene-wide association study focussing only on protein-coding genes. By looking for associations at the gene instead of variant level, we reduced the number of variables from millions to thousands. While this approach does not consider variants far outside of known gene areas (which are often implicated in genetic association studies (Schierding *et al*., 2014; Zhang and Lupski, 2015)), protein coding genes and their immediate environment, i.e. promoter and terminator regions, present a promising initial target due to being interpretable, relatively well-characterised biologically plausible factors of disease. Lastly, instead of traditional univariate association tests that work based on effect size (the estimation of which is highly dependent on sample size (Visscher *et al*., 2014)), we used a statistical method which allowed us to prioritise genes based on the stability of association (Meinshausen and Bühlmann, 2010). This increased our power to detect small but consistent effects and thus reduced false positive findings.

Using this approach, we investigated the genetic background of three ADHD-related brain networks (Duan *et al*., 2018; Liu *et al*., 2020). We identified a set of 10 genes based on the stability of association with these three networks, examined their expression in the brain, and tested their joint association with ADHD symptoms. With this we aimed to contribute to a growing understanding of the biological pathways from genes via brain to behaviour in ADHD.

## Methods

### Participants

Our sample consisted of n=899 Dutch participants from the NeuroIMAGE (von Rhein *et al*., 2015) and the International Multicentre persistent ADHD genetics Collaboration (IMpACT) (Franke *et al*., 2010) projects. It includes both adults (n=429) and adolescents (n=470) with ADHD and healthy controls. Ethical approval was obtained from the National Institute of Health registered ethical review boards and written, informed consent was obtained from every participant. Individuals with IQ<70, epilepsy, schizophrenia, austism spectrum disorder, and neurological/genetic disorders that might mimic ADHD were excluded. A majority of the sample consisted of related siblings: there were 265 singleton participants (29%), 226 sibling duos (50%), 46 trios (15%), and 11 quartets (5%). From each participant genome-wide genetic data, brain imaging, and extensive phenotyping was collected, described below and summarised in Table 1.

**Table 1:** Sample description. N: sample size. Age: mean (standard deviation) in years. ADHD symptoms: mean (standard deviation) of ADHD symptom count. ADHD≤2: Number of participants with no more than two ADHD symptoms per domain. ADHD≥7: Number of participants with seven or more ADHD symptoms per domain.

| | N | %Female | Age | Mean ADHD symptoms | N with ADHD≤2 symptoms | N with ADHD≥7 symptoms |
| --- | --- | --- | --- | --- | --- | --- |
| Adults | 429 | 53% | 25.4 (9.4) | 6.7 (6.1) | 168 | 202 |
| Adolescents | 470 | 44% | 14.6 (2.2) | 6.4 (6.4) | 210 | 220 |

#### ADHD-related measures

ADHD symptoms were assessed in accordance with DSM-IV criteria. NeuroIMAGE assessed ADHD symptoms using a diagnostic algorithm that included the Conner’s rating scale (Conners, 1997); IMPaCT used the ADHD Rating Scale (DuPaul *et al*., 1998). Both studies assessed attention/working memory using the Wechsler Adult Intelligence Scale Digit Span test (Wechsler *et al*., 2000), a task in which subjects have to repeat a series of numbers forwards or backwards. A full description of assessment procedures and data collection can be found elsewhere (von Rhein *et al*., 2015; Franke *et al*., 2010).

### Neuroimaging

Previous work in the adult portion of our sample identified five GM brain networks that were associated with attention tasks or ADHD symptoms (Duan *et al*., 2018). Three of these networks were later replicated in an adolescent sample (article in preprint (Liu *et al*., 2020)). The complete neuroimaging procedures are described elsewhere (Duan *et al*., 2018; Liu *et al*., 2020). In short, MRI neuroimaging was performed at two scan sites using three 1.5T scanners. Both manual and statistical quality checks were performed to exclude low quality scans. Scans were normalised, smoothed, and corrected for age, sex, and scanner. GM brain networks were estimated in adults only using independent component analysis. A brain network can be interpreted as a collection of brain regions in which voxels express similar GM patterns across participants. Network loadings, i.e. individual contribution of a network to total GM variation, were calculated for each participant. Network loadings in adolescents were calculated using projected adult networks (Liu *et al*., 2020).

As mentioned, three GM brain networks were associated with ADHD-related measures in both adolescents and adults. Two networks were associated with digit span performance and one with inattention symptom count. The three networks considered in this study are

#2. the inferior frontal gyrus, which was associated with both maximum forward and backward digit span performance;

#3. the superior and middle frontal gyri, which were positively associated with maximum backward digit span performance; and

#4. the cerebellar tonsil and culmen, which were negatively associated with inattention symptom count.

The brain networks are indicated as network #2, #3, and #4, following the numbering in the earlier papers (Liu *et al*., 2020; Duan *et al*., 2018). Brain networks #1 and #5 were not considered in this paper because of inconsistent results between adults and adolescents. After correction for family relations and 20 genetic principal components, there were no significant differences in network loadings between adults and adolescents (Welch’s t-test; p=.60, p=.92, p=.76 respectively). These data are shown in Figure 1.

**Figure 1:**
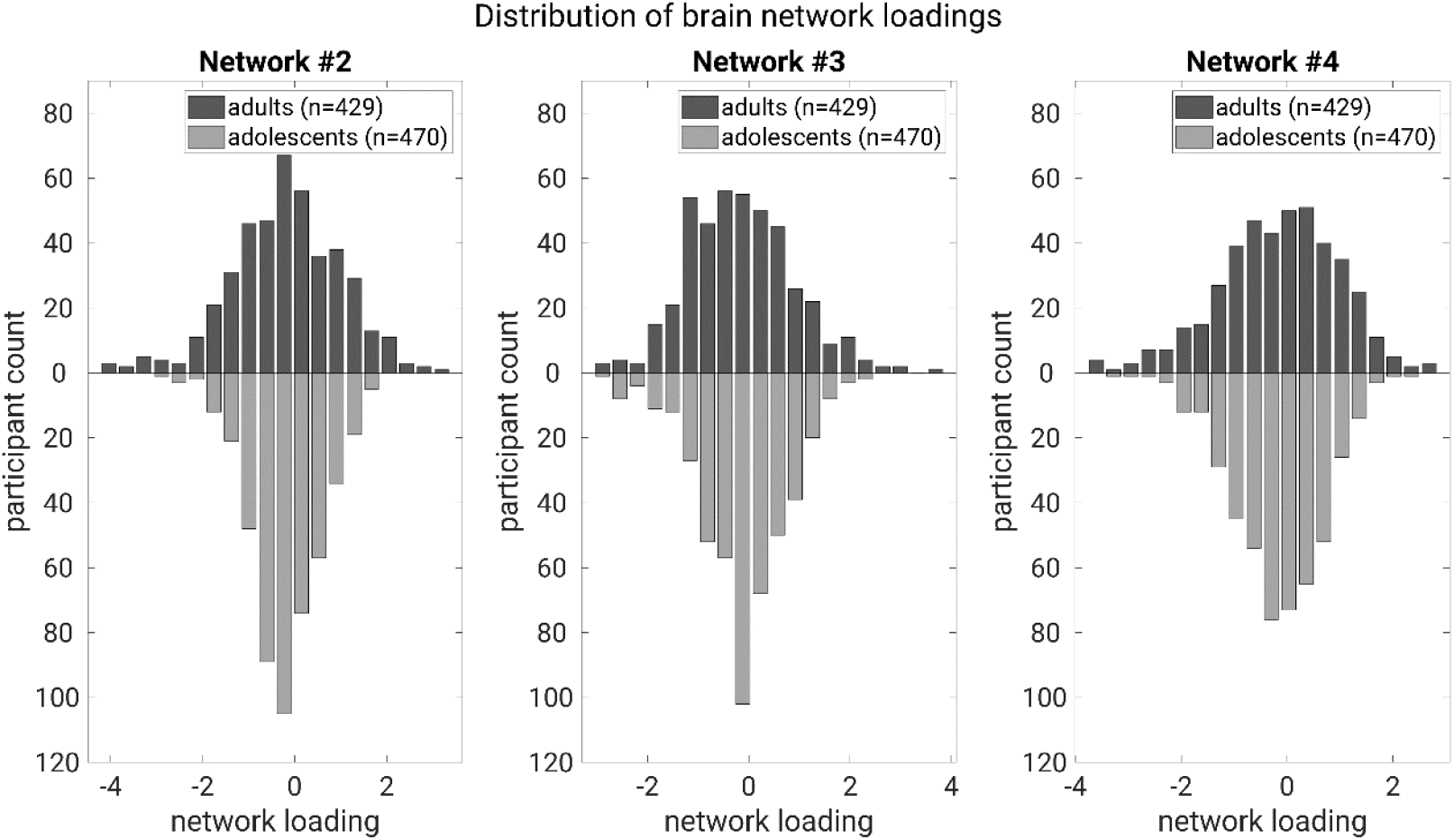
Raw (non-normalised) network loadings for adults (top) and adolescents (bottom) brain networks. Data have been corrected for age, sex, scan site, family relation, and 20 genetic principal components. No significant differences appear between adults and adolescents.

The network loadings are not independent: networks #2 and #3 are moderately correlated with r=.58 (.54-0.62 95% CI, p<1e-10), whereas loadings of both networks #2 and #4 as well as loadings of networks #3 and #4 are weakly correlated with r=.15 (.08–.21, p<1e-10), and r=.18 (.11–.24, p<1e-10) respectively. Each network is also negatively correlated with ADHD symptom count with correlations of r=-.06 (-.13–.00, p=.05), r=-.08 (-.15–-.01, p=.01), and r=-.11 (-.18–-.05, p<.001) respectively.

### Genetics

For NeuroIMAGE, genome-wide genotyping was performed using the Perlegen and HumanCytoSNP-12 genotyping platforms. For IMpACT, it was performed using the PsychChip platform. Variant-level quality control included >95% call rate, >1% minor allele frequency (MAF), and a Hardy-Weinberg Equilibrium p-value >1e-6. Participants were excluded if the individual call rate was below 95% (K. Brookes *et al*., 2006; Neale *et al*., 2010; von Rhein *et al*., 2015). Imputation was performed using the ENIGMA Consortium imputation protocol (Thompson *et al*., 2014), which in short consists of imputation using the 1000 Genomes reference population (1000 Genomes Project Consortium *et al*., 2015) and Minimac imputation software (Fuchsberger *et al*., 2015; Howie *et al*., 2012).

Genetic variants were mapped to gene level using the quantitative genetic scoring (QGS) method ((Schoenmacker *et al*.); under review). This method calculates phenotype-agnostic scores for selected genetic regions representing a relative difference from a reference population. QGS values for 12,407 protein coding genes were calculated using variants (MAF >1%) with the European section of 1000 Genomes as a reference population (Consortium *et al*., 2010; 1000 Genomes Project Consortium *et al*., 2015). Only protein coding genes were considered to limit the number of variables in our analyses. Gencode release 29 (Frankish *et al*., 2019) was used for genes and gene locations and a flanking region of 10kb before and 35kb after every gene was included. QGS values were normalised before analysis.

### Statistical analyses

Three independent stability selection analyses were performed, one for each brain network. Stability selection was performed using the randomised lasso procedure (Meinshausen and Bühlmann, 2010). One random lasso permutation step consisted of the following procedure. First, a random subsample of size n/2 was selected (without repeats). The next steps were performed on this subsample. Second, every predictor (i.e. protein coding gene QGS value) was multiplied by a random weight between .5-1. This random weight is a hyperparameter and its purpose is to better detect the stability of correlated variables. Third, corrected network loadings were regressed against all genes in a lasso-penalised multiple regression with a series of *m* decreasing penalisation strengths. This means that progressively more predictors are selected by the lasso model. The result is a 12,407 by *m* binary matrix containing whether a particular gene was selected at a particular penalisation strength.

This procedure was performed for 5000 subsamples for each network. Afterwards, the 5000 binary matrices were summed together and divided by the number of permutations to produce a stability path per gene, consisting of a stability score (i.e. the selection probability) for each penalisation strength.

The penalisation strengths were determined at the start of an analysis and not varied. They consisted of a geometric series beginning with a value just large enough so that no genes are selected and decrease until (on average) about 500 genes are selected. This value of 500 genes corresponds to an upper bound of the per-family error rate *E(V)* of 100. This upper limit on the error rate was based on the expected number of non-zero variables for a given penalisation strength (Meinshausen and Bühlmann, 2010).

We focused on genes reaching a stability score of .5, or in other words, genes that were selected in more than half of the random subsamples. Any gene with a stability >.5 we call consistently associated or “consistent” for short. These consistent genes represent a prioritised selection of the associations with the highest stability.

For all consistent genes, univariate association to the relevant brain network was calculated using linear regression. These associations are not independent from the stability selection and are expected to be strong. Similarly, for all consistent genes univariate associations with ADHD symptoms were calculated. These associations are mostly independent from the stability selection (but not completely due to the -.11 to -.06 correlation between brain networks and ADHD symptom count) and are expected to be weak.

Association of the set of consistent genes with ADHD symptoms was tested using a permutation approach where a set of *s* genes (where *s* is the number of consistent genes) were randomly selected and tested for association with ADHD symptoms. This was repeated 10,000 times. In addition, a polygenic risk score (PRS; (Purcell *et al*., 2009)) approach was used to test the proportion of variance in ADHD symptoms explained using 5, 10, 50, 100, 250, and 500 genes with the highest stability. To this end, the sum of these genes multiplied by their respective brain network association effect size was calculated. Then this sum score was associated to ADHD symptom count. Significance, again, was tested using a randomised gene permutation approach and corrected for multiple testing.

Brain expression of the consistent genes was tested using Human Protein Atlas data (available at proteinatlas.org) (Uhlén *et al*., 2015). The Human Protein Atlas classifies protein-coding genes into several categories: elevated expression in the brain (compared to non-brain tissue); non-specific expression including brain; elevated non-brain expression; and no expression in brain at all. A binomial test was used to determine whether a significant number of consistent genes fell into the first two categories (elevated brain expression or non-specific brain expression).

## Results

### Stability

In total ten consistent genes were identified, meaning that these had a selection probability ≥ 0.5. The results are shown in Table 2. The overall most consistent gene was RELL1 for brain network #4, being selected nearly 60% of the time. As a result, it was also significantly associated with this network (β=-.14, 95%CI (-.20–-.07), p=3.5e-5). Note that overall, eight out of ten regression coefficients for the consistent genes were negative, including RELL1.

**Table 2:**
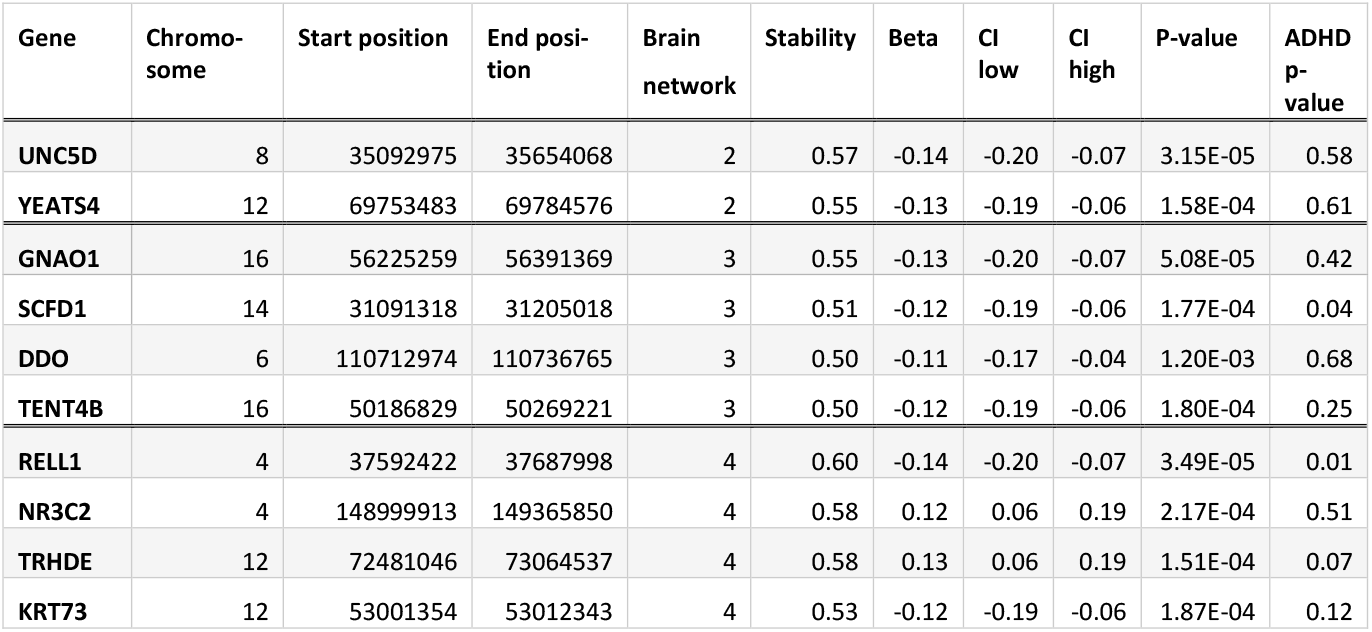
Consistently associated genes. Below are the ten genes that had a stability ≥ 0.5. The “Beta” column represents the univariate regression coefficient for the gene with the relevant brain network. The 95% confidence interval for the beta is given, as well as the uncorrected p-value. The “ADHD p-value” column represents the uncorrected p-value for the univariate association to ADHD symptom count.

For network #2, the most consistent gene was UNC5D, being selected almost 57% of the time. For network #3 this was GNAO1 with almost 55%. The full stability paths are shown in Figure 2. The stability for all 12,406 genes for all three networks can be found in supplementary Table 1. This table also contains the Ensembl stable gene IDs.

**Figure 2:**
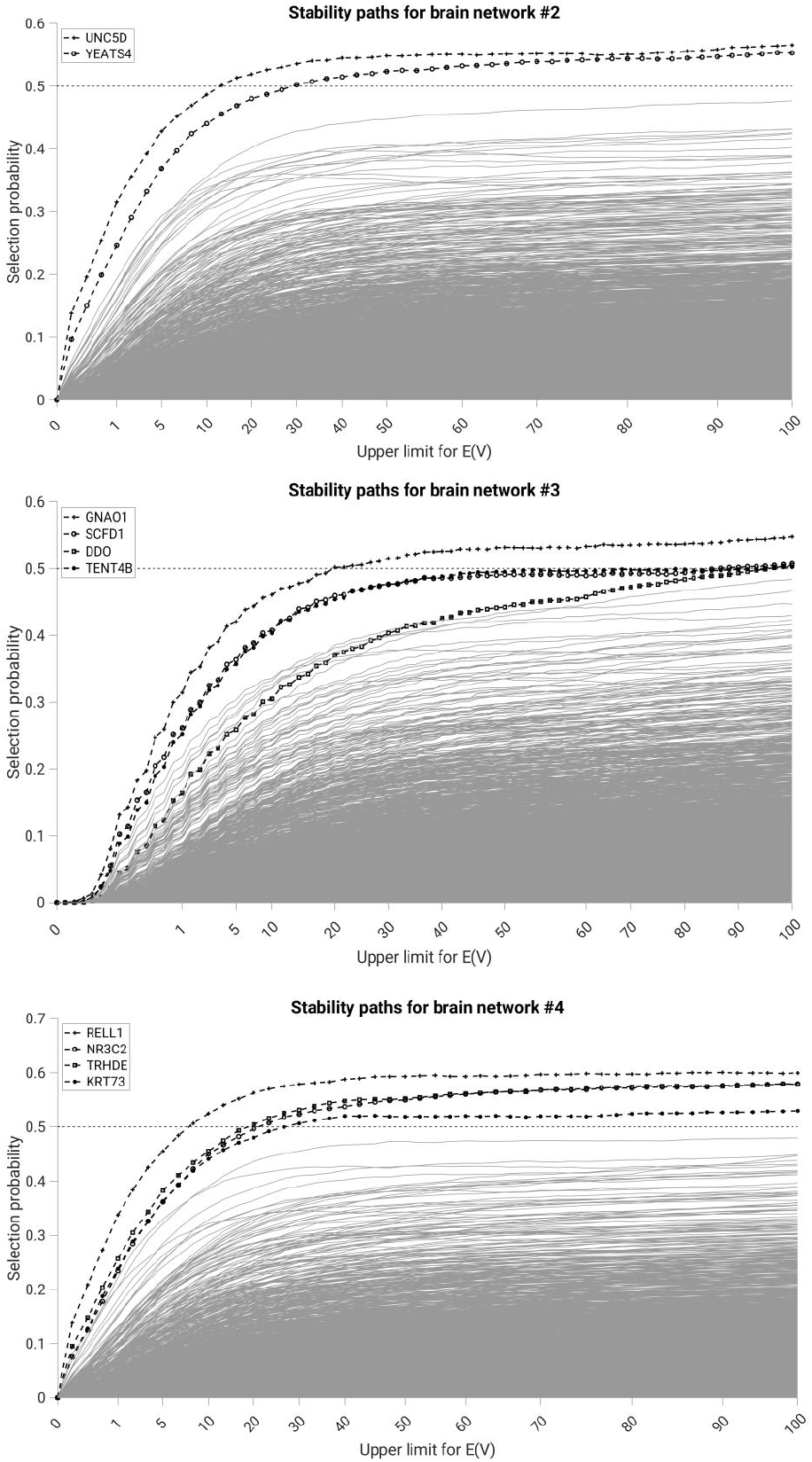
(a) Stability paths for network #2; (b) Stability paths for network #3. (c) Stability paths for network #3. The x-axis shows the upper limit for the expected number of false positives E(V).

### Expression in brain

Out of the ten consistent genes, three genes (GNAO1, TRHDE, and UNC5D) had elevated brain expression. An additional six were non-specifically expressed in the brain. This means that brain-expressed genes were significantly over-represented in our ten consistent genes (p=.03). The only consistent gene not expressed in the brain was KRT73.

### Association with ADHD symptoms

In addition to the highest stability score, RELL1 also had the lowest univariate association p-value with ADHD symptoms in our sample with p=0.01(see Table 2). The ten consistent genes together were significantly associated with ADHD symptoms when compared to ten randomly selected genes (p=.03). In addition, the PRS results showed nominally significant associations with ADHD symptoms for all networks. The genes of network #4 were significantly associated with ADHD symptoms with a maximum R^2^=.014 (p=.001) using 50 genes and the genes of network #3 were significantly associated with ADHD symptoms with a maximum R^2^=.010 (p=.009) using 500 genes. The full results are shown in Figure 3.

**Figure 3:**
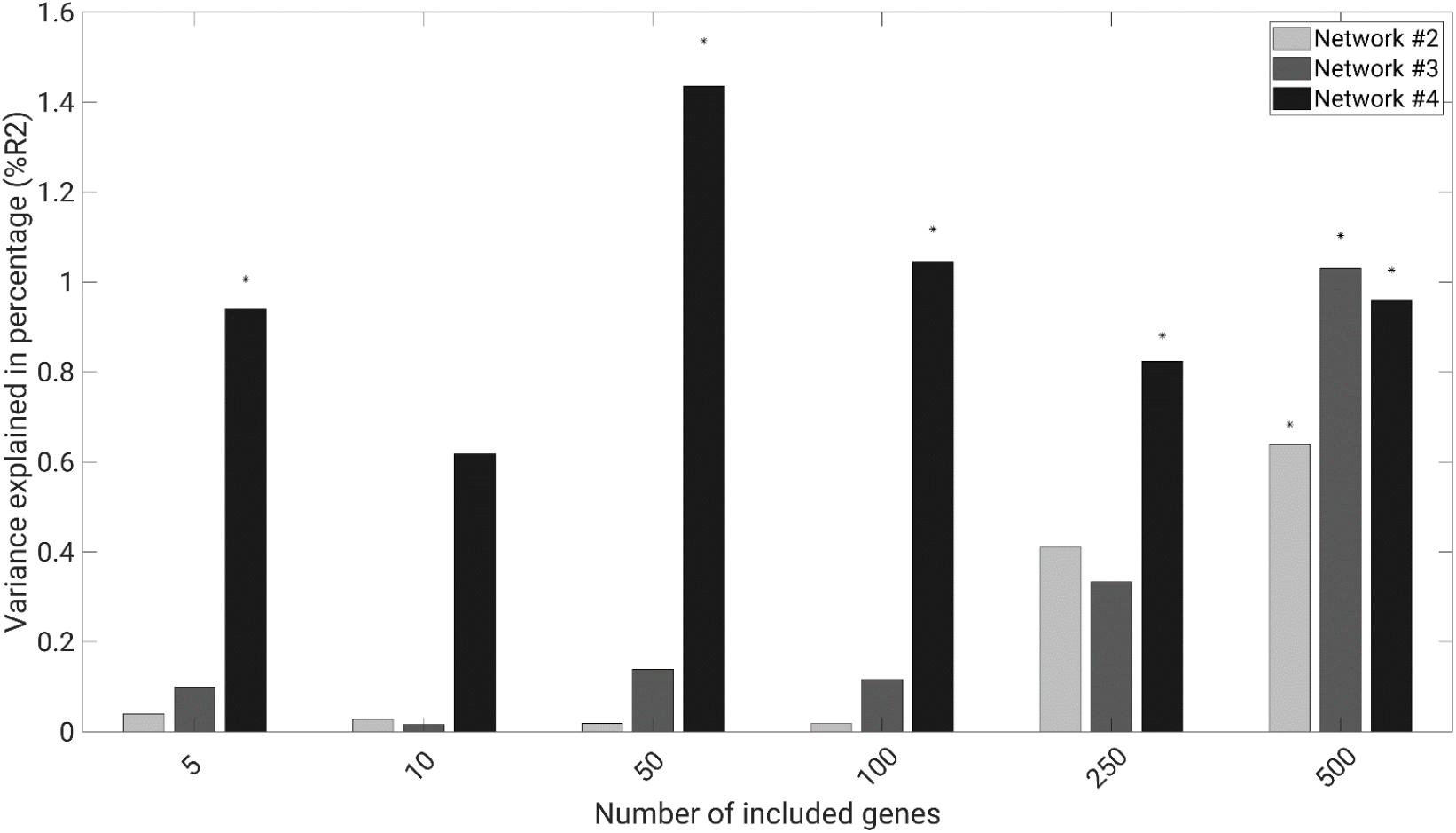
Results of ADHD symptom PRS using the top genes from stability selection. Bars with an asterisk are nominally significant at p≤.05.

## Discussion

We examined the genetic background of three ADHD-related grey matter brain networks using a resampling-based stability selection approach. We found ten consistently related genes to be associated with those networks, nine of which were expressed in the brain. Additionally, this set of ten genes was significantly associated with ADHD symptoms both as a gene set and as a variable in PRS.

The most consistent results were found for network #4, the cerebellar tonsil and culmen. The RELT-Like Protein 1 (RELL1; stability score 0.60) gene showed both the most consistent association with a brain network and the most significant association with ADHD symptoms. RELL1 encodes a receptor expressed in lymphoid tissues which is involved in apoptosis (Moua *et al*., 2017). RELL1 is unspecifically expressed in the brain and a common variant in this gene has been tentatively implicated in general cognitive function (rs111283315; p=1.86e-6) (Locke *et al*., 2015). The second-most consistent gene for this network was Nuclear Receptor Subfamily 3 Group C Member 2 (NR3C2; stability score 0.58), which encodes the mineralocorticoid receptor, a binding point for corticosteroid hormones (ter Heegde *et al*., 2015). Reduced mineralocorticoid receptor expression in the brain has been associated with schizophrenia, bipolar disorder, depression, and suicide (Klok *et al*., 2011; Medina *et al*., 2013; Qi *et al*., 2013; Xing *et al*., 2004; Young *et al*., 1998) and various classes of antidepressants increase expression in rats (Yau *et al*., 1995; Reul *et al*., 1994; Seckl and Fink, 1992; Bjartmar *et al*., 2000). The gene itself has been associated with ADHD and the HPA-axis (Kortmann *et al*., 2013). Signifcantly, the gene was also predictive of ADHD in conjunction with psychosocial stress in a partially overlapping sample (Van Der Meer *et al*., 2017). The third-most consistent gene was Thyrotropin Releasing Hormone Degrading Enzyme (TRHDE; stability score 0.58), which (as the name suggests) inactivates thyrotropin releasing hormone. The TRHDE gene overlaps with Tryptophan Hydroxylase 2 (TPH2), sharing 159 variants in our data. TPH2 variants have been associated with ADHD (Waider *et al*., 2011; K Brookes *et al*., 2006; Baehne *et al*., 2009; Sheehan *et al*., 2005) and – although conflicting findings also exist (Geissler *et al*., 2017; Sheehan *et al*., 2007; Johansson *et al*., 2010) – a meta-analysis concluded that TPH2 variants may affect executive functioning and inattention as well as other neuropsychiatric disorders (Ottenhof *et al*., 2018). In a recent study, TRHDE has been associated with a slower increase in cerebellum white matter (Brouwer *et al*., 2020).

Two consistent genes were associated with network #2 consisting of the inferior frontal gyrus. The overall fourth-most consistent gene Unc-5 Netrin Receptor D (UNC5D; stability score 0.57) encodes a receptor for netrin NTN4, which plays a role in neurite outgrowth in the cortex of rodents (Hayano *et al*., 2014). In humans, UNC5D has been associated with neuronal function (Srikanth *et al*., 2018; Sasaki *et al*., 2008) and implicated in ASD (Walker and Scherer, 2013; Hussman *et al*., 2011; Gamsiz *et al*., 2013). The YEATS Domain Containing 4 (YEATS4; stability score 0.55) gene encodes glioma amplified sequence 41, a highly preserved transcription factor with the highest expression in the brain (Munnia *et al*., 2001), which is essential for cell viability and growth (Park and Roeder, 2006). YEATS4 has been associated with depression and its treatment response (Yamagata *et al*., 2017; Amare *et al*., 2018).

The remaining four consistent genes were associated with network #3, the superior and middle frontal gyri. The most consistent gene was G Protein Subunit Alpha O1 (GNAO1; stability score 0.55), which encodes for a heterotrimeric G protein. GNAO1 function contributes significantly to synaptic neurotransmission and neurodevelopment and is associated with neurological disorders such as epilepsy, movement disorders, and developmental delay (Feng *et al*., 2018, 2019; Larrivee *et al*., 2020; Saitsu *et al*., 2016; Danti *et al*., 2017). In addition, variants in GNAO1 have been associated with depression, neuroticism, circadian rhythm, and cognitive performance (Baselmans *et al*., 2019; Jansen *et al*., 2019; Lee *et al*., 2018). The second-most consistent gene Sec1 Family Domain Containing 1 (SCFD1; stability score 0.51) has been prioritized as a causal gene for ADHD in different brain regions including cerebellum and hippocampus (Fahira *et al*., 2019) and has been associated with Alzheimer’s disease (Stamati *et al*., 2019). The D-Aspartate Oxidase (DDO; stability score 0.50) gene plays a crucial role in the central nervous and neuroendocrine systems (Katane *et al*., 2015) and has been associated with depression (Howard *et al*., 2018). Lastly, the Terminal Nucleotidyltransferase 4B (TENT4B; stability score 0.50) gene is involved in RNA metabolism and has been associated with schizophrenia (Goes *et al*., 2015).

In short, three of our ten selected genes have previously been implicated in ADHD. Other associated phenotypes include depression (4/10), cognitive performance (2/10), and schizophrenia (2/10). The strong link between our consistent genes and neuropsychiatric phenotypes suggests that the stability selection prioritisation method is able to identify promising candidates for ADHD imaging genetics. This is further reinforced by the significant over-representation of brain-expressed genes in our consistent set, as well as the small (i.e. variance explained of around 1%) but significant association with ADHD symptoms in our sample.

This significant association with ADHD symptom count could in part be explained by the correlation (-.11 < r < -.06) between network loadings and ADHD symptoms. We did not statistically correct for this possibility, because we did not expect genes to have a fully independent effect on behaviour. In fact, a post-hoc mediation analysis (data not presented) showed that the effect of the selected genes on ADHD is fully mediated by the brain networks.

Eight out of ten genetic associations with the brain networks were negative, meaning that (because of the negative correlation between networks and ADHD) their association with ADHD was positive. This imbalance in direction of effect was predicted our previous work (Schoenmacker *et al*.) and may be explained by the fact that genetic rarity – encoded by QGS as difference from the general population – is often associated with risk. Recent findings suggest that evolutionary pressure against ADHD may exist in modern environments (Esteller-Cucala *et al*., 2020) and our results lend modest support to the idea that genetic variation is more often a risk for ADHD than a protective factor.

In summary, we identified consistent associations between ten genes and ADHD-related brain networks and discussed their function and association, but we did not investigate underlying causative biological mechanisms. Employing purely statistical approach, our findings represent a list of candidates and should be seen as a starting point for further study. Further steps may include functional characterisation or bioinformatics approaches such as molecular landscape analysis. Only protein coding genes were considered, therefore, a majority of genetic variance was excluded from our analysis a priori. The currently not included genetic regions might be reconsidered for inclusion if /when larger samples become available.

By employing statistical data reduction methods in both the imaging and genetic domain and combining these with an association method based on consistency of association instead of effect size, we have created a list of ten candidate genes for ADHD imaging genetics. Three of these genes – NR3C2, TRHDE/TPH2, and SCFD1 – were previously associated with ADHD, and three have elevated expression in the brain: GNAO1, TRHDE/TPH2, and UNC5D. Our results and candidate list aid the growing understanding of the aetiology of ADHD from genes to brain to behaviour.

## Supporting information

Supplementary Table 1 (csv format)

## Data Availability

All data is available upon application though https://www.ru.nl/donders/research/research-facilities-projects/neuroimage/ and https://www.impactadhdgenomics.com/

https://www.impactadhdgenomics.com/

https://github.com/machine2learn/QGS

## Acknowledgements

This project has received funding from the European Union’s Seventh Framework Programme for research, technological development and demonstration under grant agreement no 602805 -AGGRESSOTYPE. Additional support for this work was received from the European Community’s Horizon 2020 Programme (H2020/2014 – 2020) under supported by the US. National Institutes of Health through the grant agreement no 728018 (Eat2beNICE). 1R01MH106655. The founder has no active role in this study. This work is part of the research programme Computing Time National Computing Facilities Processing Round pilots 2018 with project number 17666, which is (partly) financed by the Dutch Research Council (NWO). This work was carried out on the Dutch national e-infrastructure with the support of SURF Cooperative. This paper reflects only the authors’ views, and the European Union is not liable for any use that may be made of the information contained therein.

